# The Role of Mathematical Model in Curbing COVID-19 in Nigeria

**DOI:** 10.1101/2020.07.22.20159210

**Authors:** Chinwendu E. Madubueze, Nkiru M. Akabuike, Sambo Dachollom

**Affiliations:** Department of Mathematics/Statistics/Computer Science, University of Agriculture Makurdi, P.M.B 2373, Markurdi, Benue State, Nigeria; Department of Statistics, Federal Polytechnic, Oko, Anambra State, Nigeria; Department of Mathematics/Statistics, Akanu Ibiam Federal Polytechnic, Uwana, P.M.B 1007 Afikpo, Ebonyi State, Nigeria

**Keywords:** COVID-19, intervention measures, basic reproduction number, mass testing, social distancing, face masks, mathematical model

## Abstract

The role of mathematical models in controlling infectious diseases cannot be overemphasized. COVID-19 is a viral disease that is caused by Severe Acute Respiratory Syndrome coronavirus 2 (SARS-CoV-2) which has no approved vaccine. The available control measures are non-pharmacological interventions like wearing face masks, social distancing, and lockdown which are being advocated for by the WHO. This work assesses the impact of non-pharmaceutical control measures (social distancing and use of face-masks) and mass testing on the spread of COVID-19 in Nigeria. A community-based transmission model for COVID-19 in Nigeria is formulated with observing social distancing, wearing face masks in public and mass testing. The model is parameterized using Nigeria data on COVID-19 in Nigeria. The basic reproduction number is found to be less than unity(*R*_0_ < 1) when the compliance with intervention measures is moderate (50% ≤ *α* <70%) and the testing rate per day is moderate (0.5 ≤ *σ*_2_ < 0.7) or when the compliance with intervention measures is strict (*α* ≥ 70%) and the testing rate per day is poor (*σ*_2_ = 0.3). This implies that Nigeria will be able to halt the spread of COVID-19 under these two conditions. However, it will be easier to enforce strict compliance with intervention measures in the presence of poor testing rate due to the limited availability of testing facilities and manpower in Nigeria. Hence, this study advocates that Nigerian governments (Federal and States) should aim at achieving a testing rate of at least 0.3 per day while ensuring that all the citizens strictly comply with wearing face masks and observing social distancing in public.

## 1. Introduction

Mathematical models are logical descriptions of real-life phenomena using mathematical techniques and language. The process of developing a mathematical model is referred to as mathematical modeling [1]. Most of these models formulated are prototypes of real-life occurrences arising from science and engineering, physical system, psychology, medicine, economics and virtually all aspect of human life. These models also answer questions that emanate from changes in the behavior of the infectious organism to proffer solutions to the infections. The models use basic assumptions and mathematical approaches to determine the parameters that drive the infections and use them to proffer solutions for treatments and preventions including vaccine production [2].

Over the years, mathematical models have been tested and proven to be one of the efficient and reliable tools used in proffering control and mitigation measures to epidemics and pandemics of infectious diseases. In modeling infectious diseases, mathematical models provide practical and factual guides useful in making decisions on health policies especially concerning controlling or mitigating the virulence of infectious organisms. It can also advise health-decisions such as cost-effectiveness and optimal control of containment and intervention measures [2, 3].

The coronavirus disease 2019 simply referred to as COVID-19, is a viral disease that was first reported in Wuhan China in 2019 [4]. It is caused by Severe Acute Respiratory Syndrome coronavirus 2 (SARS-CoV-2). It has a high rate of infectivity but a low mortality rate when compared with the outbreaks of the Middle East Respiratory Syndrome (MERS) 2012 and Severe Acute Respiratory Syndrome (SARS) 2003 [5, 6]. It is known to spread through exposure to the droplets from infected persons during coughing and sneezing. As for now, Covid-19 has neither known vaccine nor specific treatments [7]. The World Health Organization (WHO) declared COVID-19 a global pandemic in March 2020 [4].

This pandemic has disrupted activities all over the world and forced many countries to reset their economic priorities [3, 8]. Different control policies recommended by the WHO such as lockdown, quarantine, isolation, social distancing and restriction of the movement were implemented by the governments of many countries to halt the spread of COVID-19 [4, 9, 10]. Sadly, these control policies have caused economic down-turn, mortalities and morbidities, inflation, high rate of crime, lawlessness, and hunger in many countries [10, 11]. The WHO and researchers have stated that COVID-19 will remain in the world until there is a vaccine [12, 13].

Though, the disease emanated from China, it spread rapidly to Europe, America, Asia, Australia and Africa within two months. As of May 13, 2020, every African country has recorded an infection, Lesotho being the last [14]. As at 7:00 GMT on June 7, 2020, Africa had a total of 492,805 confirmed cases; 244,104 active cases; 227,204 discharge/recovered persons and 11,659 mortality due to COVID-19. South Africa is the most affected country, with Egypt and Nigeria coming second and third respectively [9, 14].

Nigeria recorded her first case on February 27, 2020, from an Italian immigrant. Within four months the country recorded as at July 7, 2020, about 29,286 confirmed cases; 16,804 active cases, 11,828 discharges and 654 deaths [9]. The government of Nigeria implemented various control policies as advocated by the WHO to halt the spread of COVID-19, yet the country is still reporting outrageous new cases daily [9]. These daily new cases have been attributed to either incompatibility of the control measures with Nigerian socioeconomic environment or error on the part of stakeholders [10, 11]. For instance, the enforcement of total or partial lockdown and shutdown of humans and the economies in the entire country especially Lagos state (the most populated and industrial state in the country), Abuja the Federal Capital Territory (the seat of administrative power of the country), Kano (the commercial centre of Northern Nigeria) and Ogun state (another industrial state in southwestern Nigeria) except for essential workers (like Food vendors, the media, healthcare workers, law enforcement agencies, etc) caused a drastic decline in the gross domestic product of the country. Other consequences include; strangulation of small and medium enterprises, increase in crime due to hardship, inflation due to scare resources available and poverty in the country. The attitude of the citizens towards the disease is frustrating the fight of COVID-19 in the country: many people still believe the disease does not kill Africans, some believe that ‘COVID-19 *is just a scam to enrich some cabal*’ [11]. Many are still organizing/attending banned public gatherings, while many are still not adhering to basic safety precautions of wearing face-mask and washing of hands with running water or use of hand sanitizers [8, 10, 11].

Few researchers have attempted to mathematically model the Nigerian scenario in a bid to control and mitigate COVID-19 in the country. The work of Madubueze et al. [3], is one of the very few mathematical models of COVID-19 that seeks to consider the optimal control analysis of some of these control/containment measures advocated by WHO. Their result revealed that the combined implementation of quarantine, isolation and public health education will greatly mitigate the virus if timely implemented. However, the study did not incorporate testing for COVID-19. Iboi et al. [8] presented a mathematical model and analysis of COVID-19 to assess the impact of face-mask in the COVID-19 spread in Nigeria. Their model also revealed that COVID-19 will be eliminated in Nigeria if the wearing of face-mask in public is complemented in Nigeria with a social distancing strategy. Although all the available data on COVID-19 in Nigeria was utilized, the study did not incorporate testing for COVID-19 in their model. Okuonghae and Omane [10] formulated a mathematical model of COVID19 to assess the impact of non-pharmacological control measures such as social distancing, use of face-mask and case detection (contact tracing and subsequent testing). Their findings revealed that with intensified disease case detection rate and at least 55% of the population adhering to the observance of social distancing and the use of face-mask in public, the disease will eventually die out. However, their model analysis utilized the data on COVID-19 cases in Lagos State alone. While Ibrahim and Ekundayo [11] outlined the importance and the need for mathematical epidemiologists in times such as this, and the general misconstrued perception of COVID-19 in Nigeria. Their findings straightened the wrong perceptions and attitudes of most Nigerians towards the disease, emphasized the need for total adherence to the control/mitigation measures recommended by the government, and also affirmed the needs for mathematical models as the most urgent and necessary tools needed to effectively control/mitigate COVID-19 in Nigeria.

From the literature the authors were able to assess, none studied the impact of the non-pharmaceutical control measures (social distancing and wearing face-masks) and testing rates on COVID-19 using all the available data in Nigeria. The study done on Lagos state may not capture the true picture of the Nigerian scenario since COVID-19 has spread to all the states in Nigeria. Therefore, this study will assess the impact of non-pharmaceutical control measures (social distancing, use of face-masks) and testing rate on the spread of COVID-19 in Nigeria.

The rest of the paper is organized as follows: Section 2, is devoted to the formulation of the model and the computation of and interpretation of the basic reproduction number are presented in Section 3. In Section 4, numerical simulations are carried out to display the effect of the testing rate and intervention measures on COVID-19 dynamics. Discussion is described in Section 5 while Section 6 is the conclusion.

## 2. Model Formulation

In this section, the formulation of a deterministic community-based transmission model for COVID-19 in Nigeria is presented. The total population, *N*(*t*), at time, *t*, is divided into subpopulations; Susceptible population, *S*(*t*), Exposed population, *E*(*t*), Infected population, *I*(*t*), Isolated Infected individuals through mass testing, *I_J_*(*t*), Infected individuals that escaped mass testing, *I_NT_*(*t*), Hospitalized infectious population, *H*(*t*), and Recovered individuals, *R*(*t*).

The human population at any given time, *t*, is given by

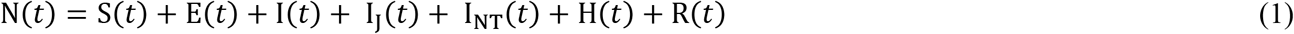

The entire population is assumed to be constant, hence there is no immigration or emigration due to border closure, birth and natural death within the pandemic is negligible. The Susceptible population, *S*(*t*), exits the compartment due to the force of infection at the rate, *λ*.

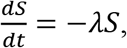

where 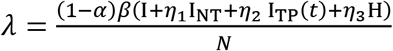 with *α* ∊ (0,1) as the intervention measures which reduce the transmission between the susceptible individuals and infected individuals. These intervention measures include wearing a face mask and social distancing. The parameters, *η*_1_, *η*_2_, *η*_3_ are the modification parameters for the subpopulation, I_NT_(*t*), I_TP_(*t*), and H(t) respectively with *η*_1_ > *η*_2_,*η*_3_. The parameters, *η*_2_, *η*_3_ are associated with the hygiene consciousness of the infected individuals that know their status that is detected and the hospitalized/isolated individuals.

The Exposed compartment gains population from the infected susceptible individuals at the force of infection rate, *λ*. They exit the exposed population after the incubation period of the virus, which is 5 – 6 days and become infected at a rate, *σ*. This is given by

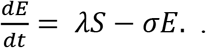

The Infected compartment, *I*(*t*), gains population from the Exposed individuals at the rate, *σ*. Mandatory mass testing is introduced in the Infected population as a diagnostic measure. This is to fish out infected individuals in the population especially those asymptomatic infected that are not aware of their status. A proportion, *p* ∊ [0,1] of the infected individuals know their status through mass testing a rate, *σ*_2_, and are isolated while the proportion, (1 − *p*), of the infected population escaped the mass testing at a rate, *σ*_1_. This yields

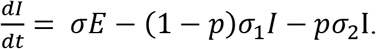

On the other hand, the Infected individuals that escaped mass testing, *I_NT_*(*t*), gains population from the proportion, (1 − *p*), of the infected individuals, at a rate, *σ*_1_. Some of them (symptomatic infected) becomes hospitalized at the progression rate, *k*, while some die of the virus at a rate, *λ*_1_. The asymptomatic infected individuals among them recover through a boost in their body immunity at a rate, *γ*_1_.

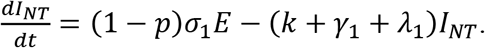

The Isolated Infected compartment, *I_J_*(*t*), gains population from the proportion, *p*, of the infected individuals, at a testing rate, *σ*_2_. They are isolated at home or a facility for observation of symptoms. The asymptomatic among them recovered at the rate, *γ*_2_, while the symptomatic are hospitalized at the rate, *θ*, with the probability, *ρ*, of developing symptoms. This gives

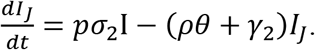

The Hospitalized infectious compartment, *H*(*t*), gains population from the infected individuals that miss mass testing at a rate, *k*, and the Isolated infected individuals that become symptomatic at a rate, *θ*, and *ρ*, the probability of developing symptoms. Some people in the hospitalized compartment recovers at the rate, *γ*_3_, while some died of the disease at the rate, *λ*_2_.

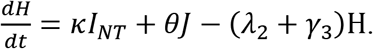

The Recovered compartment, *R*(*t*), gains population from the infectious individuals that escape testing, the isolated infected individuals and the hospitalized individuals at the respective rates, *γ*_1_, *γ*_2_, and *γ*_3_.

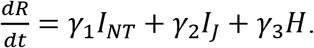

The following are the assumptions of the model.

i. The model assumes that COVID-19 is an epidemic, hence demographic processes of birth and natural death are negligible during the epidemic.
ii. The model assumes a constant population during the epidemic since borders have been closed (i.e. no immigration or emigration), hence the model is a community-based transmission model of COVID-19 in Nigeria.
iii. It is assumed that recovered individuals developed permanent immunity to COVID-19 since it has not been proven that recovered individuals become susceptible again. It is based on literature [3, 8].
iv. Infection is acquired via direct contact and inhalation or swallowing of infectious human fluid droplets.
v. Rodent infection is not considered due to the reality that present infections that are bedeviling the country are a secondary infection and do not originate in Nigeria.
vi. For an individual to become infectious, he/she must pass through the latent stage.

The schematic diagram of the transmission dynamics of the COVID-19 model is shown in Figure 1 below.

**Figure 1.**
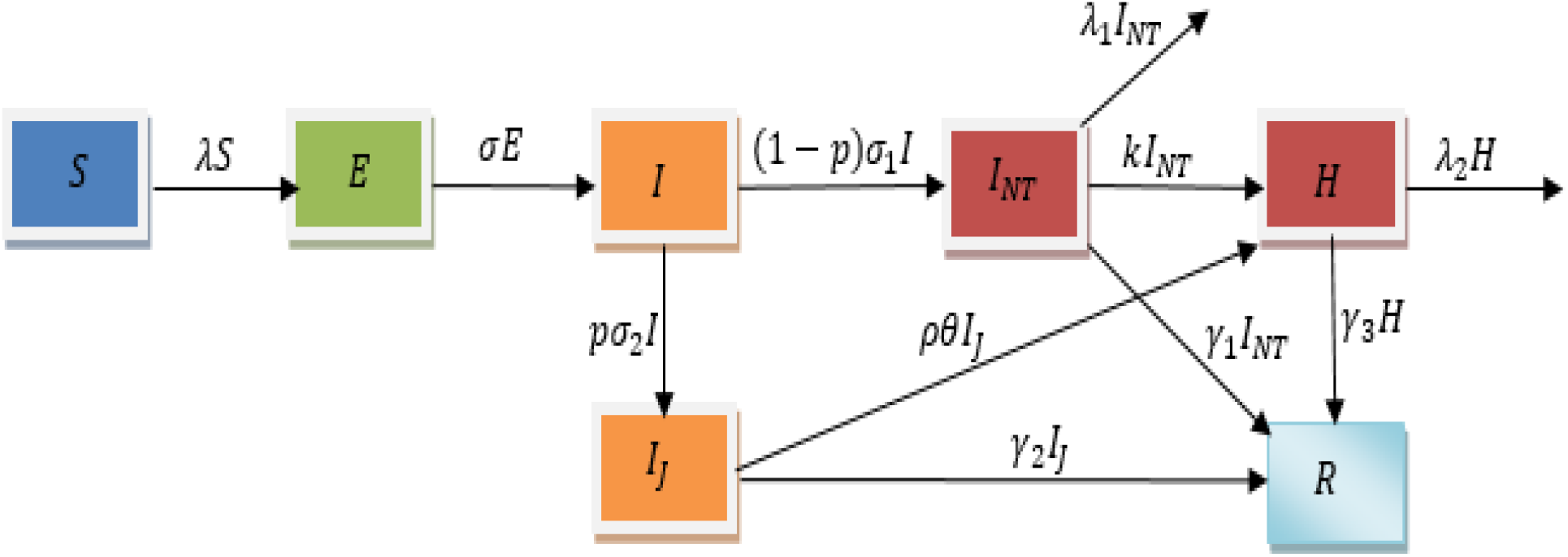
The systematic diagram of the COVID-19 model

With the assumptions of the model and the schematic diagram, the model equation is derived as

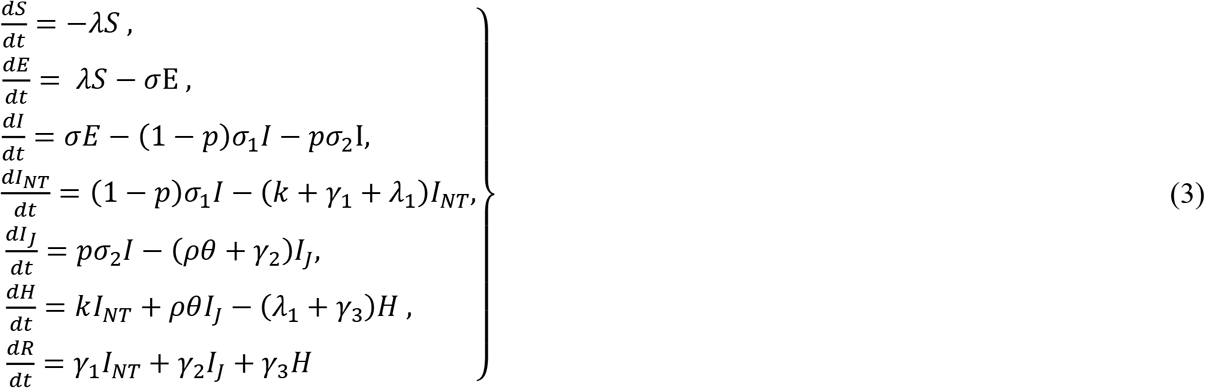

with S(0) > 0, *E*(0) ≥ 0, *Q*(0) ≥ 0, *I*(0) ≥ 0, *H*(0) ≥ 0, *R*(0) ≥ 0 as the initial conditions.

**Table 1.** **Description of Parameters of the Model**

| Variables | Description |
| --- | --- |
| $\sigma_1$ | The rate which the Infected population miss mass testing |
| $\sigma_2$ | The testing rate for the infected population |
| $\beta$ | Transmission rate |
| $k$ | The Hospitalization rate for infected individuals not-tested, |
| $\theta$ | The Hospitalization rate for isolated infected individuals, |
| $p$ | The proportion of infected individuals that participant in mass testing |
| $\rho$ | Probability of isolated infected individuals to develop symptoms |
| $\gamma_1$ | The recovery rate for infected individuals that escaped testing. |
| $\gamma_2$ | The recovery rate for isolated infected individuals |
| $\gamma_3$ | The recovery rate for hospitalized individuals |
| $\lambda_1$ | Disease induced death rate for infectious individuals that escape testing |
| $\lambda_2$ | Disease induced death rate for Hospitalized individuals |
| $\alpha$ | Intervention measures parameter |

## 3. Basic Reproduction Number

The basic reproduction number, *R*_0_, in an epidemiological study shows how transmissible or infectious an infection is. It is the average number of new infections an infected person can infect in an infective period. From the value of the reproduction number, we can predict if an infection will spread in exponential progression, die off after some time or remain constant without any further spread. When *R*_0_ < 1, the infection will die off as each infected will infect less than one person in the infective period. When *R*_0_ = 1, the infection becomes endemic and stays with each infected person infecting one new person. When *R*_0_ > 1, an infection spreads and the number of infected people will grow in an exponential proportion which will eventually lead to a pandemic as is seen in Nigeria and most of the world now for COVID-19 cases.

The basic reproduction number is computed at the disease-free equilibrium state using the next-generation approach by Van den Driessche and Watmough [15]. It is defined as the maximum eigenvalue of the matrix, *FV*^−1^ where matrices, *F* and *V* are the Jacobian matrices for new infections and the movement in or out of the infected populations by other means respectively. This is evaluated at disease-free equilibrium state, *E*_0_ = (*S*(0), 0,0, 0, 0, 0, 0,0) and the infected populations are *E*,*I*,*I_NT_*, *I_J_*, *H*. These matrices are given as

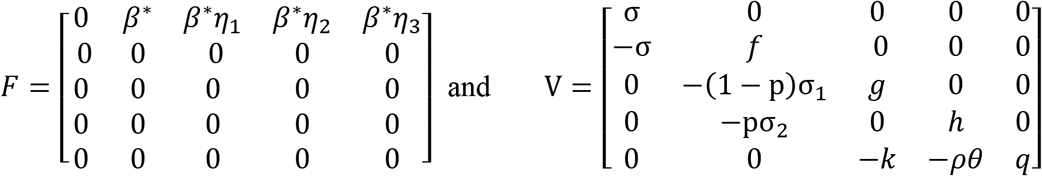

where *β\** = (1 − *α*_1_)*β*, *f* = (1 − *p*)*σ*_1_ + *pσ*_2_, *g* = *k* + *γ*_1_+*λ*_1_,*h* = *ρθ* + *γ*_2_, and *q* = *λ*_2_ + *γ*_3_. The basic reproduction number, *R*_0_, is given as

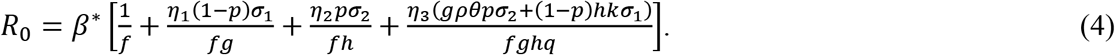

The basic reproduction number, *R*_0_, is the sum of reproduction numbers contributed by the infected populations, *I*, *I_NT_*, *I_J_*, *H*. It is represented by *R*_0_ = *R*_1_ + *R*_2_ + *R*_3_ + *R*_4_

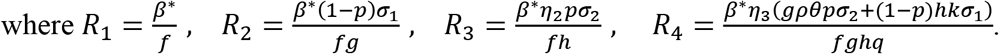

Epidemiological Interpretation of R_0_

The reproduction numbers, R_1_, R_2_, R_3_, R_4_ are the reproduction numbers of individuals in *I*, I_NT_, I_TP_, H populations contributed respectively.

R_1_ means that the infected individuals in I(t) with meantime, 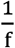, have contact with susceptible individuals, S(t) at a rate, *β\**.

R_2_ means that out of the infected individuals in I(t) with meantime, 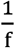, a proportion (1-p) of them miss the mass testing at a rate, σ_1_ and are represented as I_NT_ (t) and spend 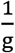 average time in I_NT_ (t) population. The I_NT_ (t) population have contact with susceptible individuals, S(t) at a rate, *β\**.

R_3_ implies that a proportion p of I(t) population with meantime, 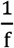, participate in the mass testing at a testing rate, σ_2_ and are represented as I_J_ population with mean-time, 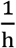. The I_J_(*t*) population have contact with susceptible individuals like health workers and family members at a rate, *β\**n_1_.

R_4_ contributes in two ways, the infected individuals that miss mass testing, 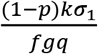 and those that participate in mass testing, 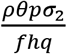. For the infected population, I_NT_ (t) that escape mass testing, they already contributed to R_2_ which is part of the reproduction number, R_4_ while the infected individuals that went in for mass testing contributed to R_3_. Some of the I_NT_ (t) and I_J_ (t) populations become hospitalized at the respective rates, k, and *θ* with the probability, *ρ*, of the I_J_ (t) population developing symptoms. They both spent a mean time of 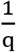 in H(t) population and have contact with susceptible individuals, S(t) at a rate, *β\**n_3_.

## 4. Numerical Simulation

Nigeria COVID-19 cases started with an imported case of a 44-year-old Italian citizen who arrived in Nigeria on 24th February 2020. He presented himself to the staff clinic of his company in Gun state on 26th February 2020 and later confirmed as the first official case of COVID-19 in Nigeria on 27th February 2020. Cumulatively, five cases were confirmed from 27th February 2020 to 18th March 2020. From 19th March 2020 to 7th June 2020, the Nigeria COVID-19 cases have risen to a five-digit number of 12801 cumulative cases [16]. On 22nd March 2020, Nigeria recorded the first death of COVID-19. The first hundred days of daily confirmed cases of Nigeria COVID-19 with their dates are displayed graphically in Figure 2 (a). The COVID-19 model of equation (3) is fit to the first hundred days of Nigeria’s cumulative cases using parameter values from the literature [3, 8]. This helps to estimate parameter values used in this work and they are presented in Table 2 with their sources. The Nigeria cumulative cases of the first hundred days with the COVID-19 model (3) are shown in Figure 2(b). The main focus of this work is to show how mass testing and intervention measures will help in curtailing the spread of COVID-19 in Nigeria. The intervention measures are face masking and social distancing. These intervention measures are already put in place by the Nigerian government. The sources in Table 2 and estimated values are the fitted parameters of the model with the cumulative cases.

**Table 2.** Parameter values for the Model

| Parameter | Value | Source | Parameter | Value | Source |
| --- | --- | --- | --- | --- | --- |
| $\beta$ | 0.487 | [8] | $\gamma_1$ | 1/7 day<sup>-1</sup> | [8] |
| $\eta_1$ | 0.2 | Estimated | $\gamma_2$ | 1/7 day<sup>-1</sup> | [8] |
| $\eta_2$ | 0.1 | [3] | $\gamma_3$ | 1/14 day<sup>-1</sup> | [8] |
| $\eta_3$ | 0.1 | [3] | $\lambda_1$ | 0.043 day<sup>-1</sup> | [8] |
| $\sigma$ | 1/5.1 day<sup>-1</sup> | [8] | $\lambda_2$ | 0.0103 day<sup>-1</sup> | Estimated |
| $\sigma_2$ | 0.061 day<sup>-1</sup> | Estimated | $\theta$ | 0.08 day<sup>-1</sup> | [8] |
| $\alpha$ | 0.5 | Estimated | $\rho$ | 0.01 | Estimated |
| $\sigma_1$ | 0.231 day<sup>-1</sup> | Estimated | $p$ | 0.1 | Estimated |

**Figure 2.**
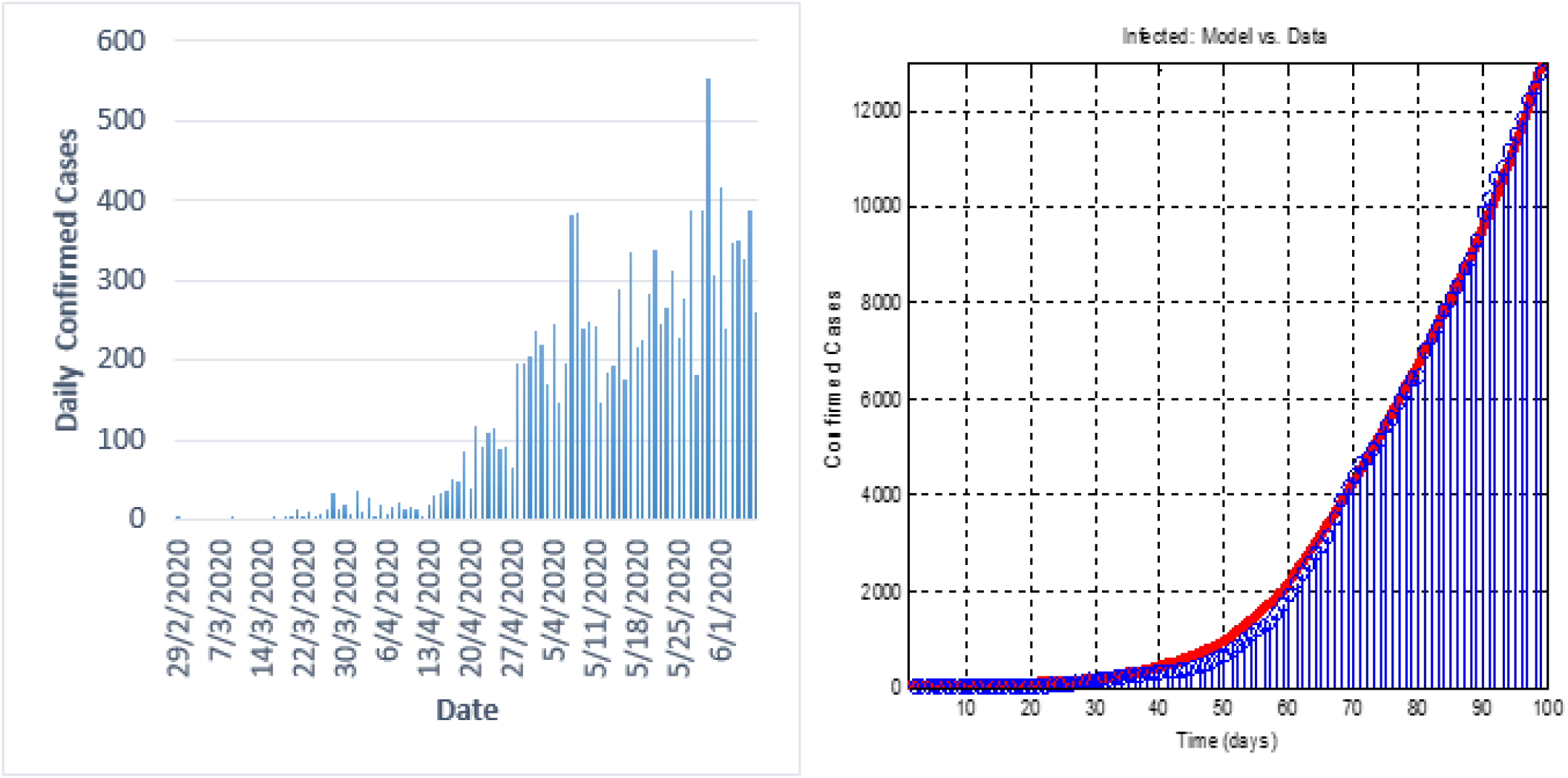
Plot displaying (a) the first hundred days cumulative confirmed cases with COVID-19 model of equation (3) (b) the first hundred Daily cases of Nigeria COVID-19.

## 5. Discussion

For the ease of understanding, we define intervention measures (face mask and social distancing) compliance as follows: i. Poor compliance: < 50%; ii. Moderate compliance: 50% ≤ *α* < 70%; iii. Strict compliance: *α* ≥ 70%. Similarly, we define the testing rate as follows: i. Poor testing rate: *σ*_2_ < 0.5; ii. Moderate testing rate: 0.5 ≤ *σ*_2_ < 0.7; iii. High testing rate: *σ*_2_ ≥ 0.7.

Using the parameter values in Table 2, the basic reproduction number (*R*_0_) for Nigeria is 1.916. This is obtained when the compliance of intervention measures is moderate (*α* = 50%) and the testing rate is poor. (*σ*_2_ = 0.061). In Figure 3, the numbers in the square boxes are the basic reproduction number values which occur at the intersection of intervention measures, *α*, and the testing rate, *σ*_2_. The compliance with intervention measures is represented on the y − axis while the testing rate is on the x − axis. The basic reproduction number is greater than unity when the compliance with intervention measures is poor (*α* < 50%) and the testing rate is high (*σ*_2_ ≥ 0.7). However, the basic reproduction number is less than unity(*R*_0_ < 1) when the compliance with intervention measures is moderate (50% ≤ *α* < 70%) and the testing rate is also moderate (0.5 ≤ *σ*_2_ < 0.7) or when the compliance with intervention measures is strict (*α* = 70%) and the testing rate is poor (*σ*_2_ = 0.3). This implies that Nigeria will be able to halt the spread of COVID-19 under these two conditions. It will be easier to enforce strict compliance with intervention measures compliance in the presence of poor testing rate caused by the limited availability of testing facilities and manpower in Nigeria. On the other hand, poor intervention measures compliance and the high testing rate will not keep the *R*_0_ less than unity. Moderate intervention measures and moderate testing rate will be needed for the *R*_0_ to be less than unity.

**Figure 3.**
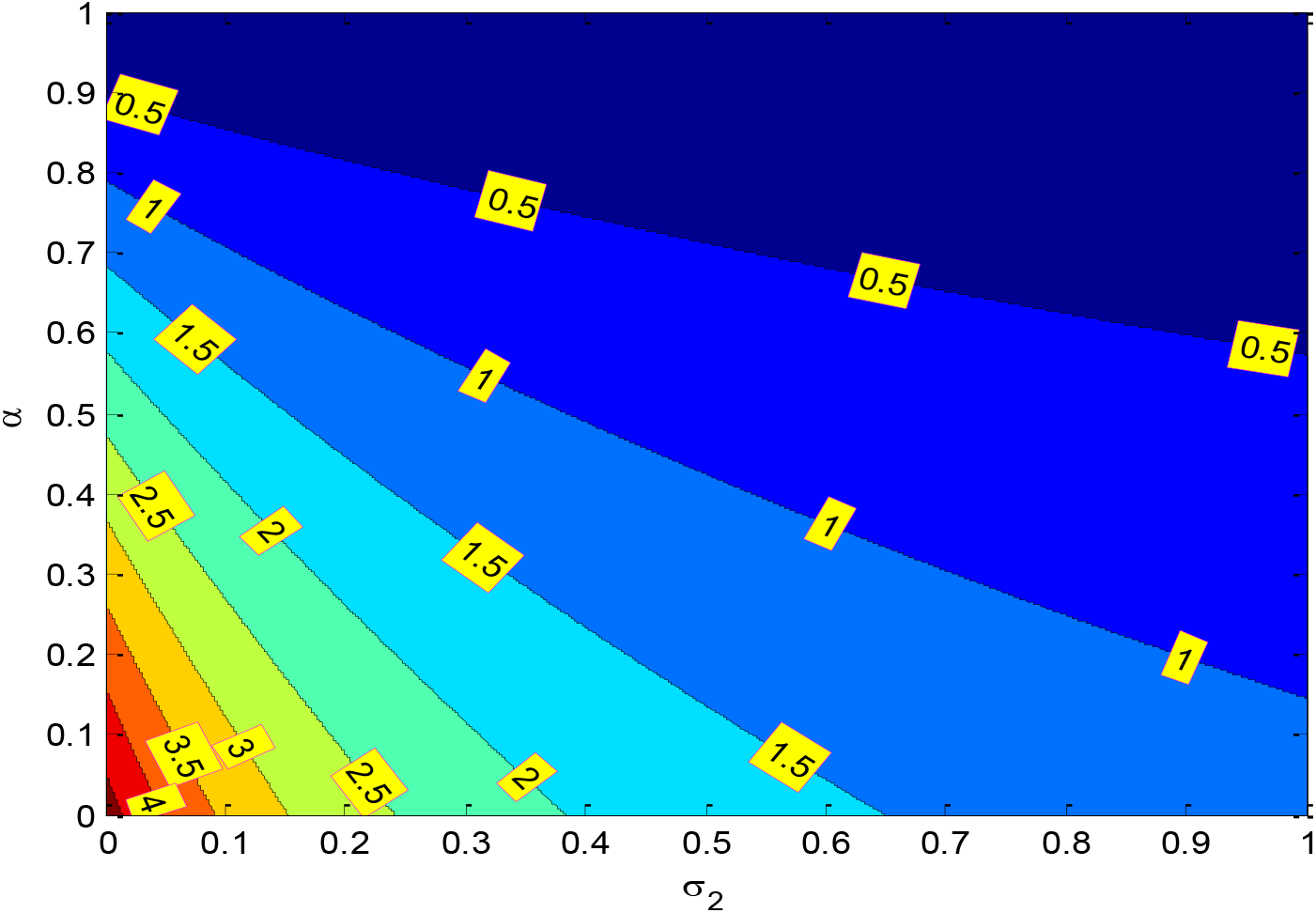
Contour plot for the basic reproduction number,(*R*_0_), of the COVID-19 model as a function of mass testing (*σ*_2_) and intervention measures, (*α*).

We observed in Figures 4(a) that when the compliance with intervention measures is moderate, the number of infected individuals in the population reduces as the testing rate increases. Also, in Figure 5(a), when the testing rate is poor (*σ*_2_ = 0.061), the number of infected individuals in the population reduces as the intervention measures compliance increases.

**Figure 4.**
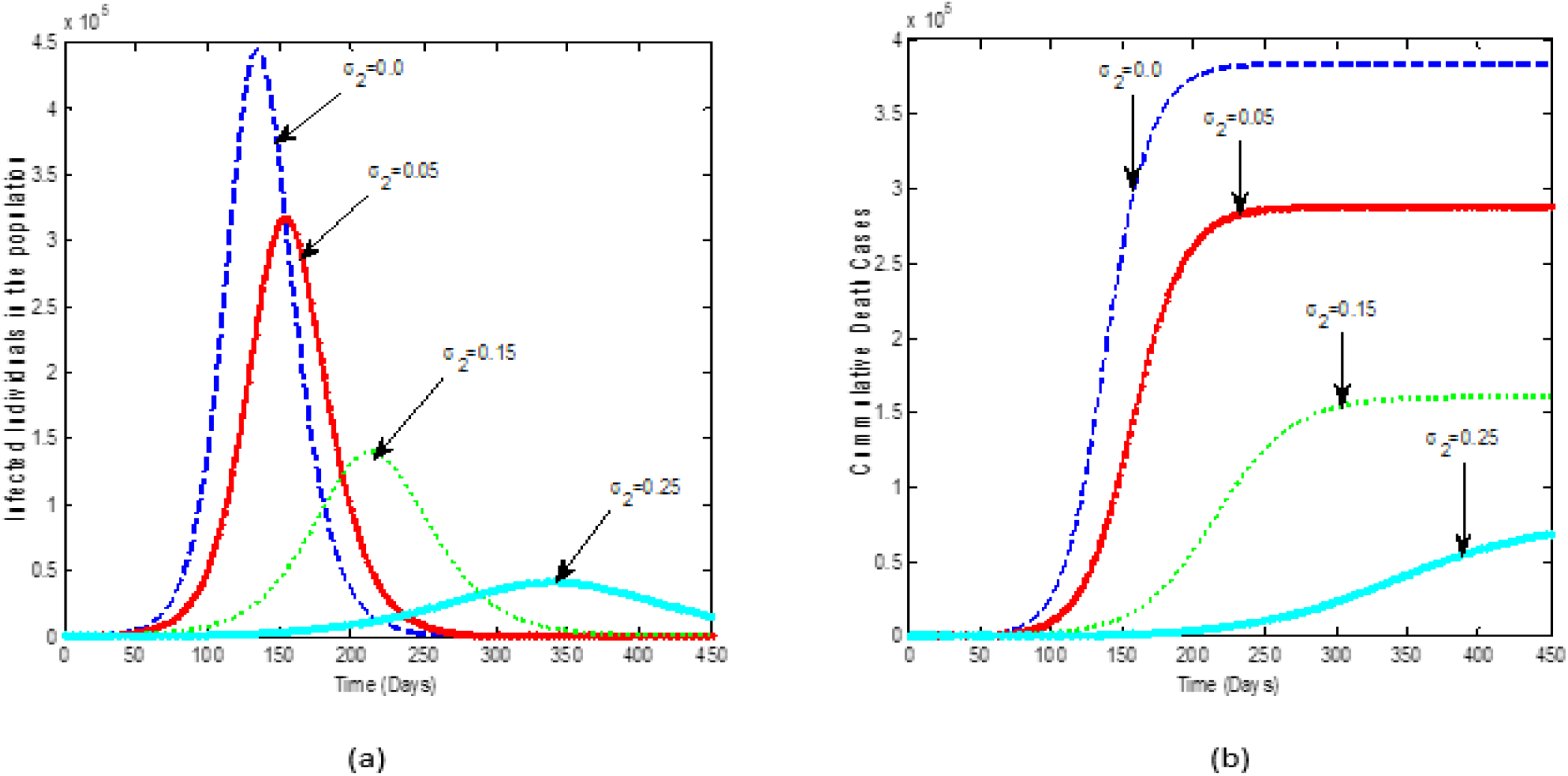
Simulation solution of the COVID-19 model of equation (3) when testing rate, *σ*_2_, is increasing in the presence of compliance of intervention measures, *α* = 50% for (a) the infected individuals in the population and (b) cumulative death cases.

**Figure 5.**
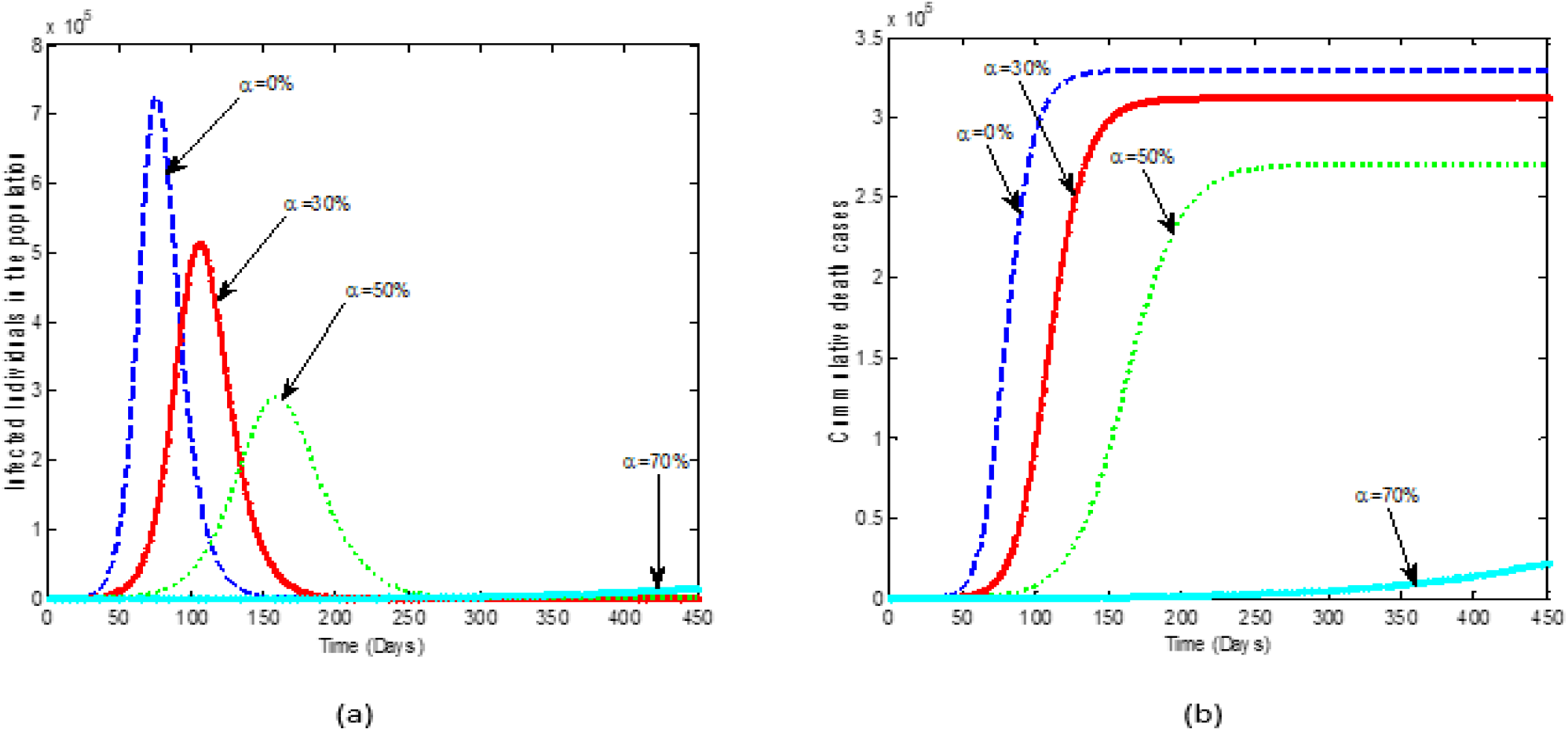
Simulation solution of the COVID-19 model of equation (3) when the compliance of intervention measures, *α*, is increasing with testing rate, *σ*_2_ = 0.061 for (a) the infected individuals in the population and (b) cumulative death cases.

Increasing both interventions reduces the number of infected individuals that could have died of COVID-19 in the country (see Figure 4(b) and 5(b)). This is expected as the infected individuals who may have contacted the virus will start treatment immediately especially the isolated asymptomatic infected individuals that have a high probability of early treatment when they develop symptoms. Death may be averted with the early commencement of proper treatment.

Figure 6(a) estimate the number of persons that will be infected when the intervention measures and the testing rates are compared at different values. For instance, when the compliance with intervention measures is poor and the testing rate is poor (*σ*_2_ = 0.3), 160,900 persons will be infected in the population. Similarly, when the compliance with intervention measures is poor and the testing rate is moderate (*σ*_2_ = 0.5), 34,460 persons will be infected. When the compliance with intervention measures is moderate and the testing rate is poor (*σ*_2_ = 0.3), 14,830 persons will also be infected. These conditions support our earlier observation that the basic reproduction number is greater than unity when the compliance with intervention measures is poor (*α* < 50%) and the testing rate is high (*σ*_2_ ≥ 0.7). This implies that COVID-19 will remain in the population unless there is an additional intervention such as vaccination.

**Figure 6.**
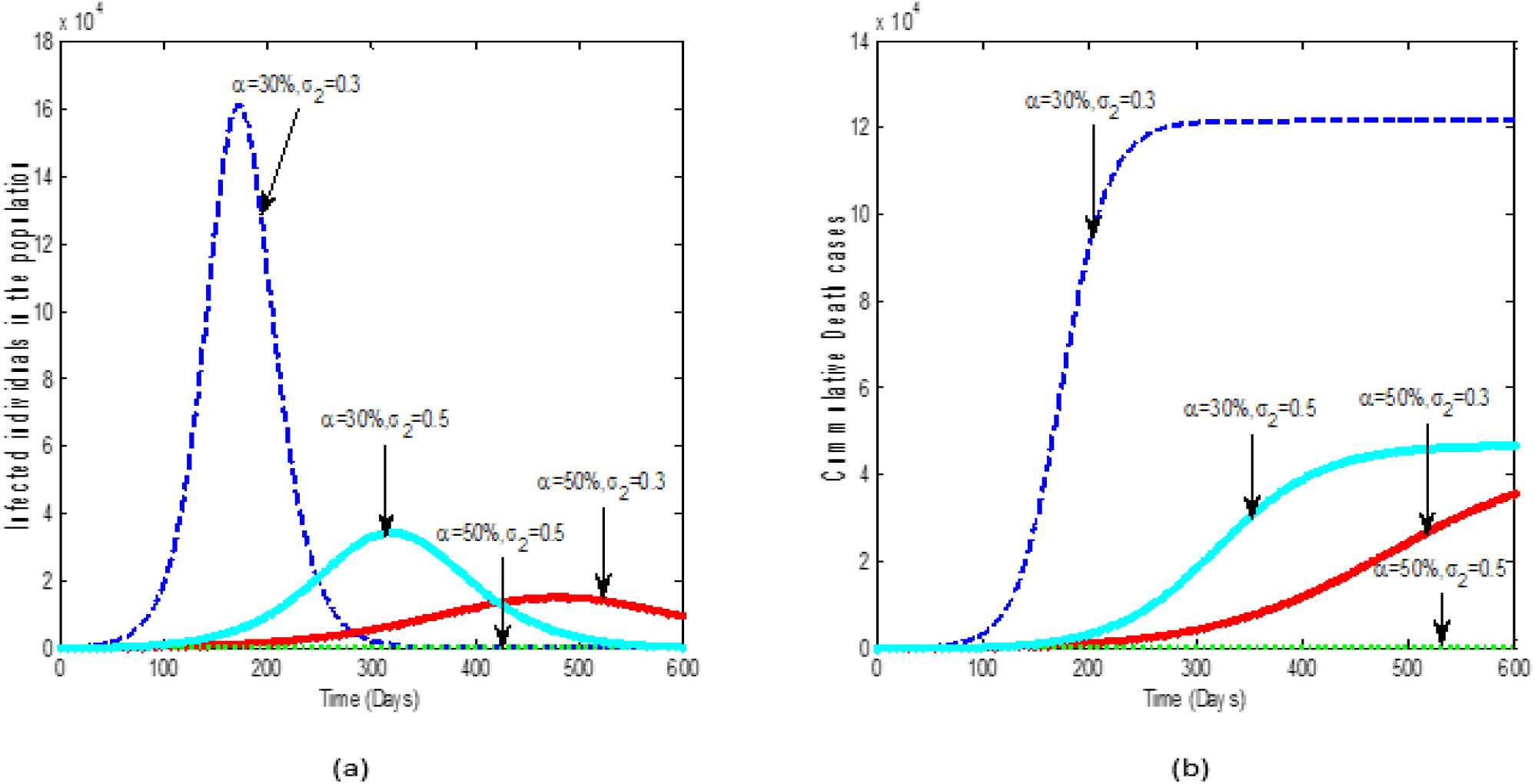
Simulation solution of the COVID-19 model of equation (3) when both the testing rate, *σ*_2_, and compliance of intervention measures, *α*, are increasing for (a) the infected individuals in the population and (b) cumulative death cases.

However, when the compliance with intervention measures is moderate and the testing rate is moderate, about 168 persons will be infected in the population. Also, when the compliance with intervention measures is strict and the testing rate is poor (*σ*_2_ = 0.3), it will result in 151 persons being infected. Any of the two scenarios will keep the basic reproduction number less than unity which implies that the chance of an infected person transmitting the infection to another person is very unlikely. These will reduce the spread of COVID-19 in Nigeria. This is similar to the finding by Okuonghae and Omame [10] who reported that the case detection rate of 0.8 per day with 55% of social distancing will eradicate COVID-19 in Lagos State Nigeria. It is also supported by the work of Iboi et al. [8] who reported that COVID-19 can be eliminated in Nigeria if social distancing and face mask compliance were at least moderate. Thus we advocate that based on limited testing facilities and lack of man-power, it will be easier to achieve strict compliance with the intervention measures even under the existing poor testing rate in Nigeria for COVID-19 to be eradicated in the population. Furthermore, Figure 6b showed that the number of dead persons will be reduced regardless of the various intervention measures compliance and testing rate combinations.

## 6. Conclusion

In conclusion, instead of the existing focus on ramping up testing rate without a set target, we advocate that Nigerian governments (Federal and States) should aim at achieving a testing rate of at least 0.3 per day while ensuring that all citizens strictly comply with social distancing and wearing of face masks. The recommended world best practices includes observing social distancing and wearing face masks.

Based on this research, it is recommended that Nigeria should target strict compliance with social distancing and wearing of face masks, and achieving at least a testing rate of 0.3 per day. This is the most plausible option with the potential to flatten the epidemiological curve of COVID-19 in Nigeria given the poor economic outlook in the country.

## Data Availability

The data for this research is within the paper

## Notes

### Competing Interest Statement

The authors have declared no competing interest.

### Clinical Trial

The research does not involved clinical trial

### Funding Statement

There is no funding for this research.

### Author Declarations

There is no IRB approval or exemption for this research.

